# G2DBridge: A Multimodal Framework Linking Genetics to Disease through Imaging Intermediates

**DOI:** 10.64898/2026.02.01.26345322

**Authors:** Okan Bilge Ozdemir, Raelynn Chen, Olivia Wu, Ruowang Li

## Abstract

Genetic-based risk prediction is becoming increasingly available for a wide range of common diseases thanks to the growth of large-scale biobanks and population-scale genetic studies. However, despite widespread availability, genetic risk prediction typically offers only modest predictive power, as complex biological processes that mediate disease manifestation are often not fully captured by genetic variation alone. In contrast, imaging-based prediction can achieve substantially higher predictive accuracy for many diseases but remains costly and unavailable for the majority of populations. Here, we present G2DBridge, a predictive machine learning framework that links genetic variation to disease risk by modeling imaging-derived phenotypes as intermediate traits. Using Alzheimer’s Disease Neuroimaging Initiative (ADNI) for training and Alzheimer’s Disease Sequencing Project (ADSP) for external validation, G2DBridge outperforms state-of-the-art polygenic risk scores in Alzheimer’s Disease (AD) prediction while relying on the same genetic data in the target data, providing additional predictive power without requiring new data modalities. The framework generalizes across cohorts, enables risk stratification with clear separation between cases and controls, and highlights biologically meaningful regions consistent with AD pathology. This approach offers a scalable strategy for applying genetically inferred imaging features in precision medicine.

## Introduction

Accurate prediction of disease risk is essential for early diagnosis, patient stratification in clinical trials, and targeted prevention strategies^1–5^. Genetic risk prediction models, such as polygenic risk scores (PRS), are widely used due to the increasing availability of large-scale genetic data from research biobanks and clinical repositories^5–9^. However, traditional PRS methods typically assume linear genetic effects^10–13^, which limit their predictive power and fail to capture individual variability, especially when key intermediate biological processes are missing from the models^14–16^.

Imaging phenotypes, derived from modalities such as neuroimaging, cardiac MRI, PET, or CT scans, have emerged as powerful biomarkers for disease prediction^17–21^. These features capture detailed structural and functional characteristics of biological systems, providing richer phenotypic information than genetic data alone^18,22^. Across diverse diseases, including neurodegenerative, cardiovascular, and metabolic disorders, imaging-based models often outperform purely genetic models in predictive accuracy^23–25^. However, the limited availability of imaging data in large-scale or routine clinical settings, due to cost, accessibility, and technical challenges^25–27^, constrains the widespread adoption of imaging-based approaches.

Existing studies have explored integrating genetic, imaging, and disease modalities to improve predictive performance^2,17,20,22,23^. Large-scale genome-wide association studies (GWAS) and derived PRS remain state-of-the-art for genetics-driven risk prediction^9,15,28^. Meanwhile, machine learning models using imaging data have demonstrated markedly superior prediction compared to genetics-only models^19,21,23,24^, but are typically trained on small imaging datasets, which restricts their broader applicability. Multi-modal integration methods that jointly model genetics and imaging have been proposed, particularly in neurodegenerative diseases^20,22,29,30^, but they generally require full access to all data modalities in the target cohort, limiting real-world usability^21^. Other approaches leverage imag-derived phenotypes (IDPs) as intermediate traits^31^ but often rely on linear PRS frameworks^32^, which lack the flexibility to capture non-linear and heterogeneous effects in imaging phenotypes. In contrast, deep neural networks (DNNs) provide a promising alternative^11–13^, as they can naturally capture complex, non-linear relationships among genetic and imaging variables without the need to specify interaction terms explicitly. DNNs have achieved state-of-the-art performance across a range of scientific domains, including image recognition, segmentation, and object detection, and are increasingly being adopted in biomedical prediction tasks^33^.

To overcome these challenges, we propose G2DBridge, a generalizable predictive modeling framework that leverages imaging-derived phenotypes as an intermediate bridge between genetic variation and disease outcomes. G2DBridge introduces several innovations compared to existing multi-modal data integration methods. First, G2DBridge learns latent imaging-derived representations using limited imaging data and transfers this knowledge to datasets where only genetic data are available, thereby improving disease prediction even when imaging is absent. Second, unlike traditional genetic risk prediction models that assume additive linear effects, G2DBridge leverages deep learning to capture non-linear and high-order genetic effects as mediated through imaging modalities on disease outcomes. By introducing imaging-derived intermediate representations, it reduces the risk of overfitting when modeling complex genetic interactions in datasets with limited imaging availability. Third, G2DBridge includes a feature extraction module that learns low-dimensional embeddings from different imaging modalities and integrates them into a unified framework. This enables multi-imaging integration while maintaining scalability to settings with incomplete imaging data.

In this study, we applied G2DBridge to Alzheimer’s disease (AD), a neurodegenerative disorder with well-established genetic and neuroimaging biomarkers^34–36^. We trained and validated our models using matched genetic data and IDPs, including FreeSurfer (FS) for brain volumes and cortical thickness^26^, Automatic Segmentation of Hippocampal Subfields (ASHS) for hippocampal regions^27^, and diffusion tensor imaging (DTI) for white matter pathways^29,30^ from the Alzheimer’s Disease Neuroimaging Initiative (ADNI). We further performed external validation using the Alzheimer’s Disease Sequencing Project (ADSP), where only genetic data were available. Our results demonstrate that G2DBridge substantially improves AD risk prediction compared to conventional PRS, when applied to the same genetic-only inputs. This integrative framework leverages limited imaging data to improve genetic risk prediction, enabling reliable and cost-effective disease risk assessment in large-scale biobanks and clinical settings.

## Results

### Overview of the proposed approach

We introduce G2DBridge, a generalizable predictive framework that links genetic variation to disease risk by modeling intermediate traits, specifically IDPs from genotype data (Figure 1). The framework leverages matched genetic, imaging, and clinical outcome data to learn cross-modal predictive relationships, effectively bridging genetics to disease through imaging-derived features.

**Figure 1.**
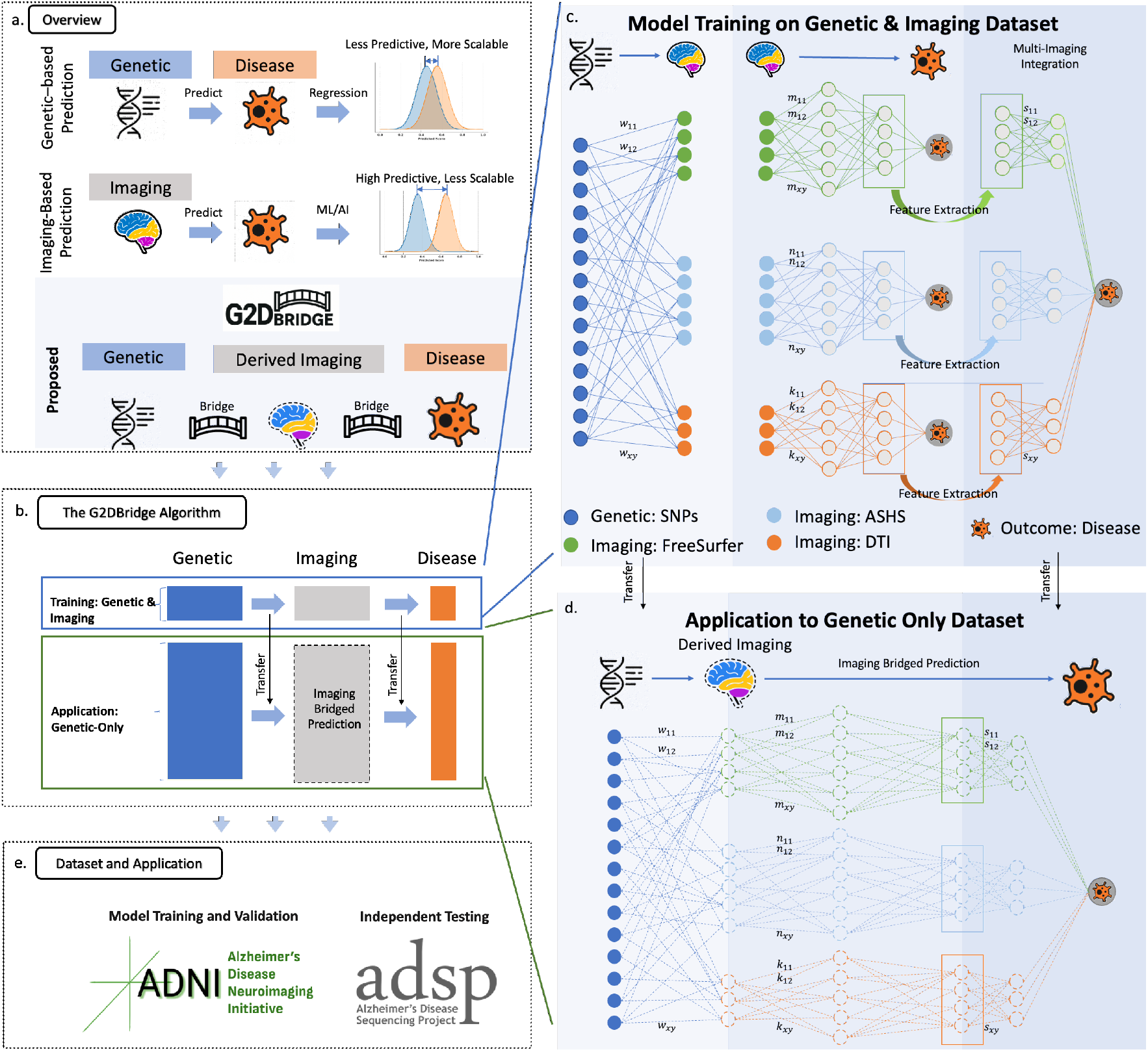
Overview of the G2DBridge framework. (a) Schematic illustration of genetics-based prediction, imaging-based prediction, and the integrative G2DBridge approach. (b) Architecture of the G2DBridge algorithm, showing training with genetic and imaging data and application with genetic data only. (c) Model training procedure using genetic and imaging data. (d) Application of the trained model to genetic-only datasets for disease prediction. (e) Datasets used for training, validation, and testing: ADNI and ADSP.

In the training phase, where all modalities are available, G2DBridge learns an intermediate representation of IDPs that connects genetic variants to disease risk. This process consists of three stages: (i) training neural networks(NN) to predict IDPs directly from genetic variants, enabling the generation of imaging-like features even in cohorts without neuroimaging data. (ii) extracting low-dimensional latent representations of each IDP from hidden layers to provide compact yet expressive embeddings. and (iii) integrating these embeddings through a downstream classification network to predict disease outcomes. In the application phase, only genetic data from the target dataset are required. The pretrained networks are transferred to infer imaging features and used for disease prediction, thereby improving risk prediction without requiring imaging data.

### AD Classification with IDPs

To evaluate the utility of IDPs for AD prediction, we compared five machine learning models (Logistic Regression, Random Forest, Lasso, XGBoost, and NN) across three IDP sources (ASHS, DTI, and FS). Figure 2-a summarizes the resulting AUC for each model-IDP combination, as well as for an integrated IDP model based on a NN.

**Figure 2.**
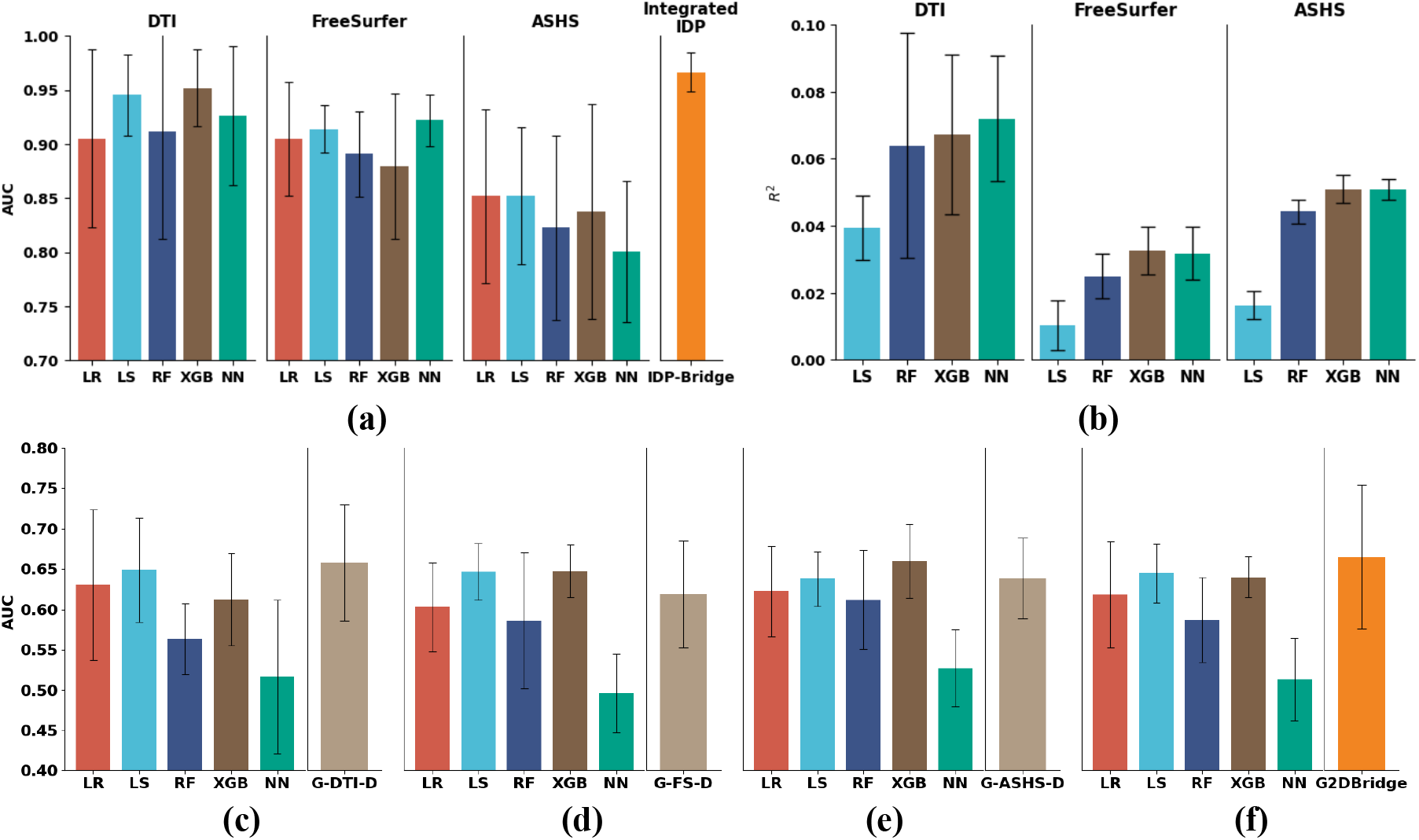
Performance of IDP based AD classification, genetic IDP prediction, and IDP-mediated G2DBridge models. **(a)** AD classification using individual and integrated IDPs. AUC performance of five machine learning models, LR, RF, Lasso (LS), XGBoost and NN, across three IDP sources (DTI, FreeSurfer, ASHS). Error bars represent standard deviations from five-fold cross-validation. The integrated IDP model (IDP-Bridge) is shown on the right. **(b)** Genetic prediction of IDPs. Explained variance (R^2^) of four machine learning models, LS, RF, XGBoost, and NN for three IDP sources (DTI, FreeSurfer, ASHS). Error bars represent standard deviations from five-fold cross-validation. **(c–f)** Comparison of individual IDP-mediated models and the G2DBridge framework. Mean AUC values with standard deviations across five-fold cross-validation. Panels show **(c)** G-DTI-D, **(d)** G-FS-D, **(e)** G-ASHS-D and **(f)** proposed G2DBridge, each benchmarked against genetic-only baselines (LR, RF, LS, XGB and NN). Error bars indicate 95% confidence intervals (CI) across folds.

In the ASHS and DTI analyses, XGBoost achieved the highest performance with AUCs of 0.838 ± 0.100 and 0.952 ± 0.036, respectively. In FreeSurfer, a neural network yielded the best performance with an AUC of 0.922 ± 0.024, followed by Lasso with 0.914 ± 0.022. Linear models, including logistic regression and Lasso, underperformed compared to the more flexible machine learning models, with 0.8%–10.5% lower AUC compared to the best model for each IDP. The multi-IDP integrated model achieved the highest accuracy with an AUC of 0.967 ± 0.018, showing that combining multiple IDPs substantially improves predictive power and that structural IDPs are robust predictors of AD diagnosis, consistent with prior studies^37–40^.

### Prediction of IDPs from Genetic Data

We next evaluated the prediction of IDPs from genetic variants by comparing four machine learning methods using explained variance (R^2^) and mean squared error (MSE). Figure 2-b summarizes the R^2^ results across models and IDPs, with corresponding MSE results in Supplementary Figure 1.

Across the three IDP sources, NN consistently provided the best balance between variance explained and low error. For DTI, NN attained the highest R^2^, 0.072 ± 0.019, while XGBoost (0.067 ± 0.024) and random forest (0.064 ± 0.034) followed with greater variability (Supplementary Figure 2). Similar patterns were observed in FreeSurfer and ASHS, where XGBoost achieved the highest R^2^ (0.033 ± 0.007 and 0.051 ± 0.004, respectively), and NN were close behind (0.032 ± 0.008 and 0.051 ± 0.003) while minimizing error. In contrast, Logistic regression and the Lasso model trailed with lower R^2^ and higher MSE. Overall, these findings establish NN as the most stable method across all three IDPs, and these results demonstrate that IDPs can both be predicted from genetic data and used to classify AD, motivating the evaluation of an integrated framework that leverages both components.

Figure 2c–f shows the performance of each IDP-mediated model compared with the genetic-only baseline. The baseline models achieved moderate performance, with mean AUCs generally in the 0.56–0.66 range, and the strongest baseline differing by IDP (DTI: LS = 0.649, FreeSurfer: XGB = 0.648, ASHS: XGB = 0.659, all-IDP baseline: LS = 0.645). In contrast, the IDP-mediated models (rightmost columns) showed mixed results across IDPs. G-DTI-D improved slightly over the best genetic-only model (AUC = 0.658 ± 0.082, +1.4% vs the best baseline), whereas G-FS-D (AUC = 0.619 ± 0.076) and G-ASHS-D (AUC = 0.638 ± 0.057) were lower than their strongest genetic-only counterparts. The corresponding area under the precision–recall curve (AUPRC) results are provided in Supplementary Figure 3.

**Figure 3.**
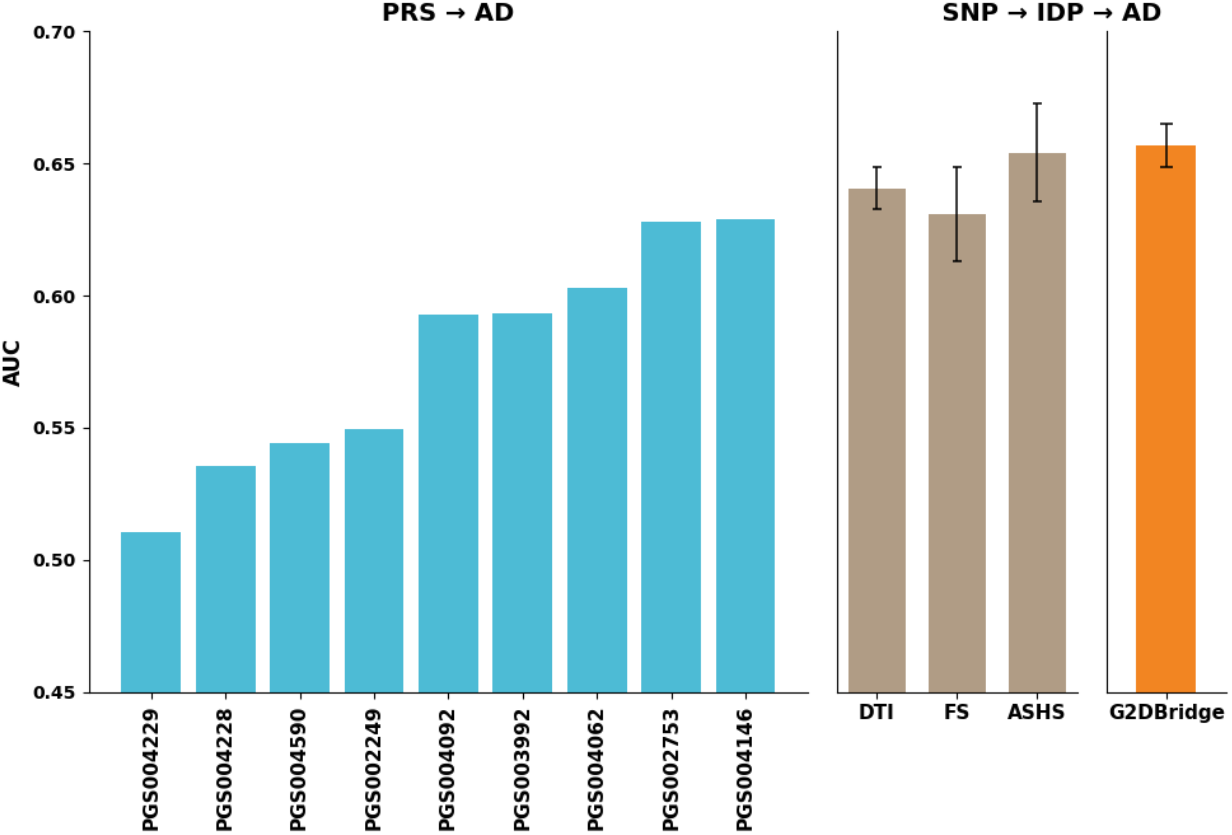
External validation on ADSP. (Left) Performance of nine polygenic risk score (PRS) models from the PGS Catalog for AD classification in ADSP; model identifiers (e.g., PGS004146) correspond to catalog entries. (Right) Performance of IDP-mediated models (DTI, FreeSurfer, ASHS) and the integrated G2DBridge model trained on ADNI and tested on ADSP. Error bars indicate 95% confidence intervals (CI) across folds.

In the multi-IDP setting, G2DBridge combined latent representations from all three IDPs using an integrated neural network architecture. This integrated model achieved an AUC of 0.665 ± 0.102, exceeding the genetic-only baselines by +7.47% (LR), +13.22% (RF), +3.12% (Lasso), +3.87% (XGB), and +29.53% (NN). Its performance was higher than G-DTI-D (AUC = 0.658), G-ASHS-D (AUC = 0.638), and G-FS-D (AUC = 0.619), indicating that integrating multiple IDPs can provide additional predictive power beyond individual IDP-mediated models in this setting.

### External Validation on ADSP Dataset

To evaluate generalizability, models trained on the ADNI dataset were externally validated on the independent ADSP cohort. As a benchmark, we first examined nine PRS models from the PGS Catalog (Figure 3, left). These models, all trained on datasets independent of ADSP, achieved AUCs ranging from 0.51 to 0.63, with the highest accuracy observed for PGS004146 (AUC = 0.629). Despite representing state-of-the-art PRS benchmarks, overall performance remained modest.

In contrast, our IDP-mediated models, which estimate imaging-derived phenotypes from genetic data before classifying AD, demonstrated improved performance (Figure 3, right). Models based on individual IDPs achieved AUCs of 0.641 (DTI), 0.631 (FreeSurfer), and 0.654 (ASHS). The corresponding AUPRC results are provided in Supplementary Figure 4.

**Figure 4.**
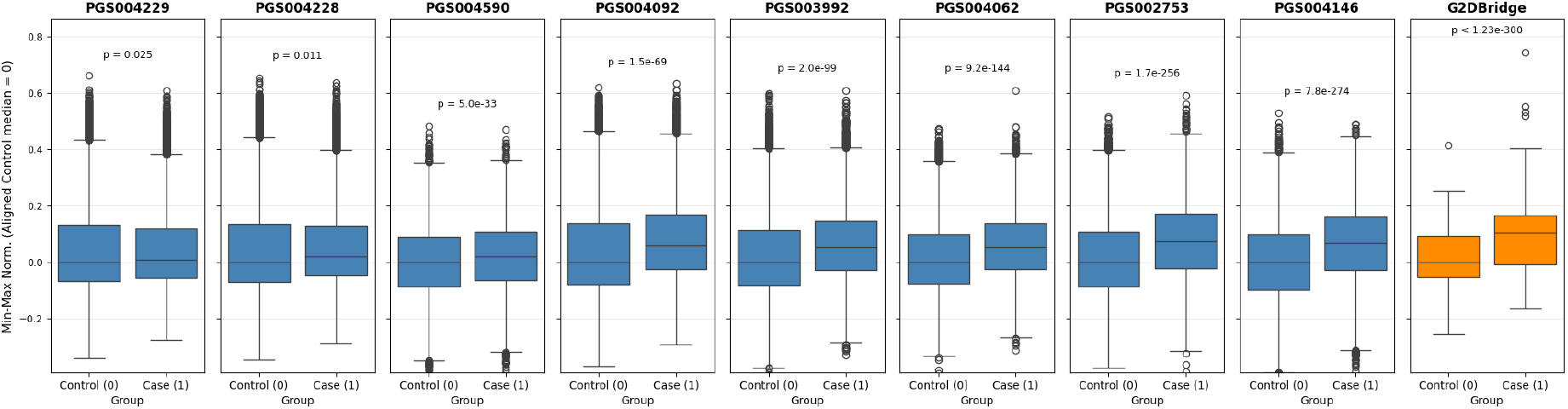
Risk stratification using PRS models and G2DBridge. Box plots showing the distribution of min–max normalized prediction scores across case and control groups. Scores are aligned such that the median of the control group is set to zero. PGS identifiers correspond to models from the PGS Catalog. Welch’s t-test was used to assess group differences; p-values are shown above each panel.

Finally, the integrated G2DBridge framework, which combines genetically predicted IDPs through a neural network, achieved the highest accuracy with an AUC of 0.657. This outperformed both the PRS baselines and individual IDP-mediated models.

Together, these results demonstrate that G2DBridge generalizes effectively to independent cohorts and highlights the value of leveraging genetically derived imaging representations for disease risk prediction.

### Risk Stratification Analysis

AUC summarizes ranking ability but does not show how prediction scores distribute between cases and controls. Risk stratification analysis addresses this by revealing group separation and areas of overlap or misclassification. We therefore examined the distributions of normalized prediction scores for each model (Figure 4). All PRS models showed statistically significant differences between cases and controls, with the strongest among them being PGS002753, PGS004062, and PGS003992. However, none matched the discriminative strength of G2DBridge, which achieved the most significant separation (Welch’s t-test, p < 1.23 × 10^−300^). These results indicate that G2DBridge captures the relationship between genetic risk and AD status more strongly than standard PRS approaches.

### Feature Attribution and Consistency Analysis

To better understand which imaging features contributed most to AD classification, we applied gradient-based attribution analysis and visualized feature importance using two complementary methods.

First, we generated beeswarm plots of the top 20 features per fold, with results from fold 1 shown in Figure 5 (a–c) (DTI, FreeSurfer, ASHS; additional folds in Supplementary Figure 5-7). These plots display subject-level signed Integrated Gradient (IG) values, where positive attributions push the model toward predicting AD and negative values toward control.

**Figure 5.**
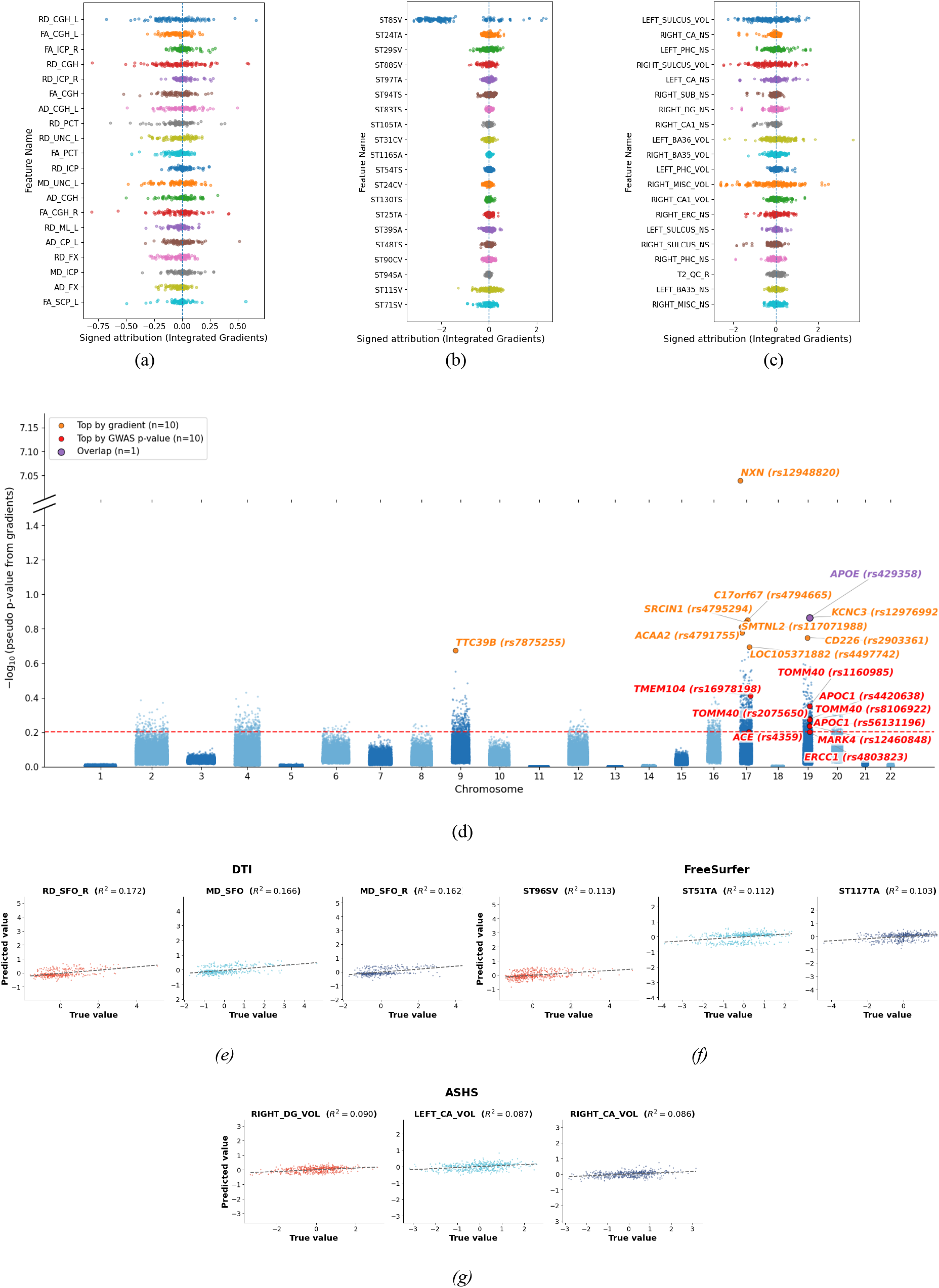
Feature attribution, SNP importance, and modality-specific predictive fidelity. **(a–c)** Beeswarm plots of subject-level signed Integrated Gradients (IG) for the top 20 features from fold 1, shown separately by IDP modality (DTI, FreeSurfer, ASHS). Points are jittered for visibility. Positive IG values push the model toward Alzheimer’s disease (AD), while negative values push away. **(d)** Manhattan plot of gradient-based SNP importance with GWAS and gene annotations. Each point is a SNP positioned by chromosome and genomic coordinate. The y-axis shows gradient-derived importance transformed as −log10 of a pseudo p-value computed from min-max normalized gradients, so larger values indicate higher model importance. Labeled SNPs are the union of two sets: orange labels mark the top gradient-ranked variants above the percentile threshold, and red labels mark variants prioritized by the lowest GWAS p-values among those same high-gradient candidates (GWAS filtering applied using the external p-value threshold). Variants meeting both criteria are highlighted as the overlap category. The red dashed horizontal line indicates the 99th-percentile cutoff (top 1%) of transformed gradient-importance scores. Gene symbols for labeled SNPs were retrieved from dbSNP and displayed next to rsIDs. The plot highlights the canonical AD locus on chr19 (APOE/TOMM40/APOC1 region) alongside additional labeled genes for follow-up. **(e–g)** True versus predicted values for the best-performing feature in each modality using the best-performing model: **(e)** DTI, **(f)** FreeSurfer, **(g)** ASHS. Each point represents one subject, and the dashed line shows the linear fit between true and predicted values.

Across modalities, distinct patterns emerged. For DTI, diffusivity metrics from the cingulum–hippocampal tract (RD_*CGH, MD_CGH*) and uncinate fasciculus (*MD_UNC_L*) were consistently among the strongest predictors, aligning with prior studies of white matter disruption in AD. In FreeSurfer, cortical and subcortical features from temporal regions (e.g., *ST119TS, ST11SV, ST8SV, ST29SV*) repeatedly ranked highly, reflecting known cortical thinning and atrophy in AD. ASHS features (e.g., *LEFT_BA36_VOL, RIGHT_BA36_VOL*) highlighted medial temporal lobe structures, including parahippocampal gyrus, entorhinal cortex, Cornu Ammonis, dentate gyrus, and subiculum, regions long established as early markers of AD-related neurodegeneration.

Together, these analyses confirm that the model relies on biologically meaningful imaging markers, with DTI capturing microstructural tract damage, FreeSurfer reflecting cortical atrophy, and ASHS identifying medial temporal lobe degeneration.

Figure 5-d summarizes SNP-level signals using a Manhattan-style layout. Each point represents a SNP positioned by chromosome and genomic coordinate, and the y-axis reports a transformed model-derived importance score computed as −log_10_(p_pseudo_), where p_pseudo_ is derived from min-max normalized gradient magnitudes. This transformation makes high-gradient SNPs visually comparable to conventional GWAS Manhattan plots. GWAS summary statistics were downloaded from the NHGRI-EBI GWAS Catalog (GCST013197), corresponding to Wightman, D. P. et al.^41^, and used only as an external reference for interpreting SNP-level signals; GWAS p-values are not plotted in this panel.

To improve readability in the presence of one extreme value, the y-axis is shown with a broken scale: most variants cluster in the lower range, whereas a single high-scoring locus is displayed in the upper panel. Labeled variants correspond to (i) the top gradient-ranked SNPs above the chosen percentile threshold and (ii) the top GWAS-supported SNPs among those same high-gradient candidates, using an external GWAS p-value filter. The union of these labeled SNPs was annotated using dbSNP to obtain mapped gene symbols, and these genes were printed to facilitate downstream manuscript edits and biological interpretation.

The labeled set includes expected Alzheimer’s disease loci in the chr19 *APOE* region (*APOE, TOMM40, APOC1, MARK4*), demonstrating that the gradient-based ranking recovers known AD-associated genomic signals. In addition, the plot highlights other genes in the labeled set (e.g., *ACE, TTC39B, KCNC3, SRCIN1, SMTNL2, TMEM104, ERCC1, NXN*), which may reflect cohort-specific effects, linkage with nearby causal variation, or model sensitivity to correlated genomic features. Overall, Fig. 5d provides a compact view of how model-derived SNP importance aligns with established AD genetics while also surfacing additional candidates for follow-up.

Figures 5e–g visualize NN predictions for each modality (DTI, FreeSurfer, ASHS). Predictions were concatenated across cross-validation folds, feature-wise *R*^2^ was computed, and the top three target features were selected for display. The true–predicted scatter plots show a clear positive, approximately linear association between ground truth and NN estimates, indicating good agreement; the dispersion around the fitted regression line reflects residual error and potential differences in calibration across targets.

## Discussion

In this study, we introduced G2DBridge, a predictive framework that links genetic variation to disease risk through IDPs as intermediate traits. By combining the broad availability of genetic data with the biological specificity of neuroimaging, G2DBridge enables scalable prediction while retaining interpretability. Although AD served as the case study because of its strong genetic and imaging biomarkers, the approach is generalizable to other neurodegenerative, cardiovascular, and psychiatric diseases where paired genetic and imaging data exist.

Our results highlight three key findings. First, IDPs are informative predictors of AD status, and integrating multiple IDPs improves classification compared with single-modality models (Figure 2-a). The multi-IDP model achieved the highest overall accuracy, showing that combining modalities provides a more complete representation of disease pathology^37–40^. Second, IDPs can be predicted from genetic data (Figure 2-b and Supplementary Figure 2), with NN outperforming other machine learning methods across modalities in both explained variance and error. Third, and most importantly, genetically predicted IDPs improve disease risk prediction compared with PRS. In ADNI internal validation, genetically inferred IDPs yielded consistently higher AUCs than genetic-only models (Figure 2-c-d-e-f). In ADSP external validation, G2DBridge outperformed multiple state-of-the-art PRS models (Figure 3), achieving superior accuracy while using the same target genetic data. This represents a free gain in predictive power. By learning intermediate imaging representations, G2DBridge extracts additional signal without requiring new data types or increased genotyping cost. From a clinical perspective, the framework also provides added value in risk stratification. Distributional analyses demonstrated significantly clearer separation between AD cases and controls for G2DBridge than for PRS models (Figure 4).

Interpretability analyses further showed that G2DBridge relies on biologically meaningful markers (Figure 5). Across modalities, distinct patterns emerged. For DTI, diffusivity metrics from the cingulum– hippocampal tract (*RD_CGH, MD_CGH*) and uncinate fasciculus (*MD_UNC_L*) were consistently among the strongest predictors, aligning with prior studies of white matter disruption in AD^42–44^. In FreeSurfer, cortical and subcortical features from temporal regions (e.g., *ST119TS, ST11SV, ST8SV, ST29SV*) repeatedly ranked highly, reflecting known cortical thinning and atrophy in AD^45–47^. ASHS features (e.g. *LEFT_BA36_VOL, RIGHT_BA36_VOL*) highlighted medial temporal lobe structures, including parahippocampal gyrus, entorhinal cortex, Cornu Ammonis, dentate gyrus, and subiculum regions, long established as early markers of AD-related neurodegeneration^48,49^.

This work has several limitations. First, the analyses pooled participants across ancestries for model training and evaluation due to limited sample sizes in several individual ancestry groups. As larger datasets become available, ancestry-specific models will likely deliver improved and more generalizable performance^49,50^. Second, we used IDPs from structural and diffusion MRI rather than raw images to reduce dimensionality and improve robustness; however, models operating directly on raw MRI may capture finer signal when adequate data, harmonization, and computation are available^50^. Future work could also include PET or functional imaging to provide more biological insight^51^. Finally, while explainable AI helped improve interpretability, it does not establish causality. Future studies could integrate causal inference methods to better understand biological mechanisms^49^.

In summary, G2DBridge generalizes across cohorts, improves prediction relative to PRS without additional data requirements, and identifies biologically meaningful imaging features. By bridging genetics, imaging, and disease outcomes, the framework offers a scalable and clinically relevant strategy for advancing genetic risk prediction and moving toward personalized medicine.

## Methods

### Data

We used two large and well-characterized cohorts: the Alzheimer’s Disease Neuroimaging Initiative (ADNI) and the Alzheimer’s Disease Sequencing Project (ADSP). All models were trained and tuned on ADNI with five-fold cross-validation, and ADSP was reserved exclusively for external validation.

#### ADNI

ADNI is a multicenter longitudinal study of older adults spanning the spectrum of AD. It provides genetic, neuroimaging, and clinical data. For this study, we constructed binary outcome models using genetic variants and IDPs. Three IDP sets were analyzed: diffusion tensor imaging (DTI), FreeSurfer morphometric measures, and Automatic Segmentation of Hippocampal Subfields (ASHS). These modalities capture complementary aspects of brain structure and were used to evaluate the contribution of single vs. multi-IDP integration. After quality control and intersection, 203,909 SNPs remained(Supplementary Table 2). IDPs were standardized using a single z-score scaler fitted on the with-genome subset for each IDP set, and the same transformation was applied consistently when generating model inputs. The analytic samples were DTI with D=265 features and N=385 participants, FreeSurfer with D=323 features and N=534 participants, and ASHS with D=35 features and N=534 participants. The intersection common to all three IDP sets contained N=355 participants. The genotype-only cohort contained N=651 individuals for the ADNI dataset.

#### ADSP

ADSP is a large consortium integrating genetic and clinical data across diverse AD studies. We used ADSP version 5 solely as an independent test set. To ensure comparability, identical quality-control procedures were applied to ADNI and ADSP. Single-nucleotide variants with minor allele frequency below 0.05 or with high missingness were removed, and analyses were restricted to variants present in both datasets. The genotype-only cohort contained N=25113 for the ADSP dataset. Individuals present in both the ADSP and ADNI datasets were removed from the ADSP dataset before performance evaluation.

#### Machine learning models

We implemented logistic regression, Lasso regression, random forest, XGBoost and feedforward neural networks to predict both continuous (IDPs) and binary (AD status) outcomes.

##### Logistic regression

Standard logistic regression (generalized linear model, no penalty) was used as a linear baseline for binary classification.

##### Lasso regression

Lasso regression (L1 penalty) was applied to enable variable selection in both regression and classification settings, particularly suitable for high-dimensional SNP data. The penalty parameter (α) was optimized using glmnet.

##### Random forest

Random forest classifiers and regressors were used to capture nonlinear relationships and assess feature importance. Hyperparameters including n_estimators, max_depth, max_features, and min_samples_split were tuned using Optuna.

##### XGBoost

Extreme Gradient Boosting (XGBoost) was employed as a powerful ensemble method that combines multiple decision trees using gradient boosting to capture complex nonlinear relationships in the data. Its regularization capabilities help prevent overfitting, making it particularly effective for high-dimensional genomic data. Hyperparameters such as n_estimators, learning_rate, max_depth, and subsample were optimized using Optuna.

##### Neural networks

Feedforward neural networks were implemented in PyTorch. Architectures varied in depth, width, activation functions, and dropout rates, all optimized with Optuna. Training used the Adam optimizer with early stopping based on validation loss.

Hyperparameter optimization was performed with five-fold cross-validation, using AUC for classification and R^2^/MSE for regression.

#### Interpretability

To improve interpretability, we used gradient-based attribution methods from Captum (PyTorch library), focusing on Integrated Gradients. These analyses estimate how much each input feature contributes to the model’s output, allowing us to quantify feature importance at both the subject and cohort levels.

### G2DBridge framework

G2DBridge is a three-stage neural network framework that links genotype data to AD risk via genetically predicted IDPs.

Stage 1: Genetics to IDPs. Three parallel multilayer perceptron map the standardized SNP vector *x* ∈ *R*^*p*^ to a predicted IDP 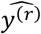, where *r* ∈ {DTI,FS,ASHS}. Each network has its own parameters θ^(*r*)^ =,*W*^(*l,r*)^*b*^(*l,r*)^/, *L*_*r*_ hidden layers and nonlinear activations θ^(*l,r*)^.

Stage 2: IDPs to AD. For each modality, a classifier predicts AD status from the standardized IDP vectors. The final hidden layer of each classifier provides an embedding *f*^(*r*)^, representing a compact modality-specific feature vector.

Stage 3: The three IDP embeddings are concatenated into a fused representation

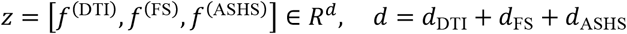

and passed through a multilayer classifier with K hidden layers. The final output is the predicted probability of AD,

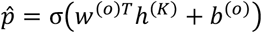

The model was implemented in PyTorch with the Adam optimizer. Hyperparameters (layer widths, depth, learning rate, dropout, activation functions) were tuned with Optuna. We used five-fold cross-validation with early stopping on validation loss. Performance was reported as mean AUC (classification) or mean R^2^ (regression) across folds.

#### Application to genetic-only data

Once trained on cohorts with paired genetic and imaging data, G2DBridge can be applied to genetic-only datasets. In this setting, the first stage of the framework predicts IDPs directly from the SNP vector, IDP 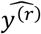. The resulting genetically inferred IDPs are passed through the second and third stages of the network (IDP→AD classifiers and multi-IDP integration) to yield AD risk probabilities.

#### Polygenic risk scores

All polygenic scores used in this work were obtained from the PGS Catalog and computed using previously published SNP effect sizes from large-scale GWAS studies. Before scoring, SNP alleles were strand-matched and harmonized with reference panels to ensure consistent allele orientation across datasets. Genome-wide PRS were then calculated across all variants passing quality control using the *plink2 --score* function.

We considered multiple Alzheimer’s disease PRS derived from large European cohorts, as well as one score trained on multi-ancestry samples. These polygenic scores were generated using various methodologies. A detailed summary of all polygenic scores, including their PGS identifiers, training sample sizes, number of variants, and methodologies, is provided in Table 1.

**Table 1.**
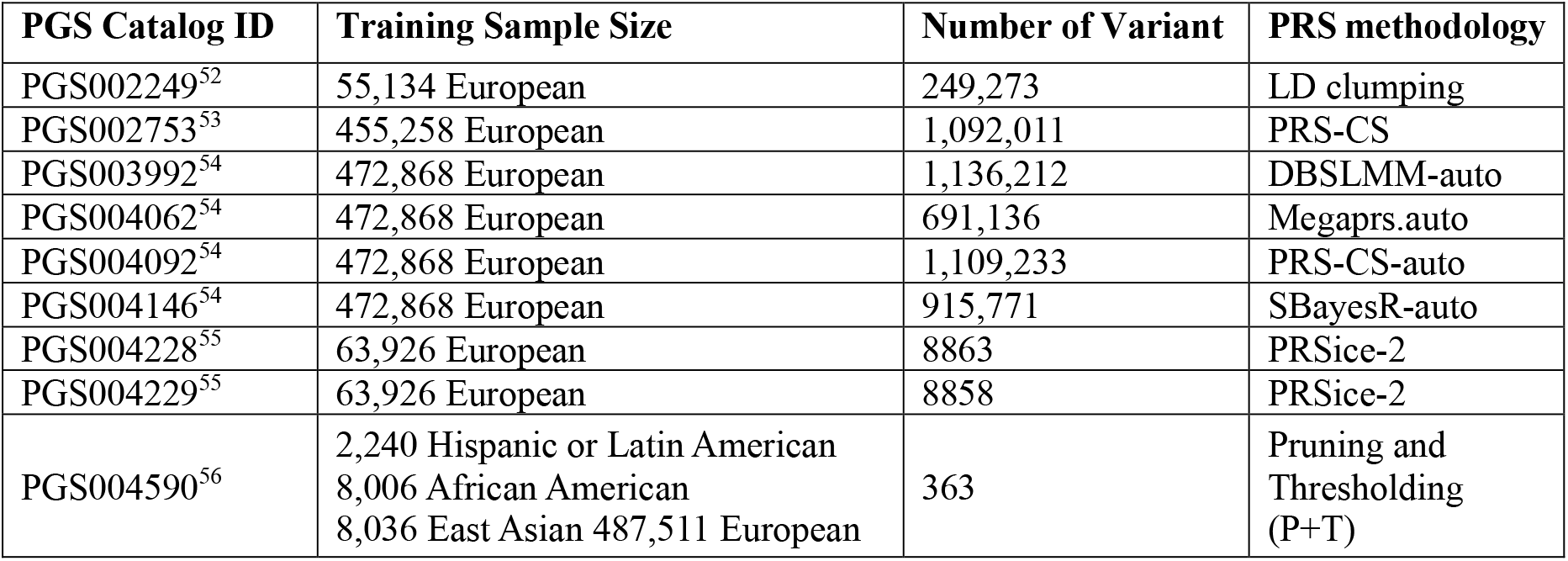
Comparison with PGS Catalog PRS.

#### Risk stratification analysis

For risk stratification, we compared the distributions of model prediction scores between AD cases and controls. Prediction scores were min–max normalized, aligning the median of controls to zero for comparability. Group differences were assessed using Welch’s t-test, and significance values were reported for each model.

## Data Availability

All data produced are available online at https://adni.loni.usc.edu/ and https://adsp.niagads.org/

## Acknowledgments

O.B.O, R.C, and R.L are supported by the Department of Computational Biomedicine, Cedars Sinai.

## Author Contributions

O.B.O. and R.L. contributed to method development, model training, and writing the paper, while R.C. and O.W. contributed to data processing.

